# Type 2 Diabetes and Heart Failure: A Comparative Analysis of Disease Trajectory and Clinical Outcomes

**DOI:** 10.64898/2026.06.22.26356266

**Authors:** Abes A. Bautista Neughebauer, Zackary Tushak, Raymond L. Benza, Deepak Talreja

## Abstract

**Background:** Approximately 40 million individuals in the US have diabetes, and 6.5 million are also afflicted with congestive heart failure (CHF). This paper outlines the natural history of CHF in T2DM and compares CHF outcomes between patients with and without T2DM.

**Methods:** We performed a retrospective analysis of prospectively collected data from 2,008 patients hospitalized for CHF exacerbation between December 2016 and June 2019. Propensity score matching was used to match diabetics and nondiabetics. Outcomes included survival and readmission rates at 28 d, 3 mo and 6 mo, as well as comparison of echocardiographic findings.

**Results:** A total of 2,008 patients were included. After matching, 492 patients were included, with 244 diabetics and 248 nondiabetics. After matching, readmission rates within 28 days (p=0.625) were not different, but there was a trend for higher readmission rates among diabetics at 3 months (29.3% vs. 21.5%, p=0.049) and 6 months (44.3% vs 35.8%, p=0.053). Echocardiographic characteristics, including LVEF (p=0.135), LV EDV (p=0.707), maximum velocity of mitral valve E wave (p=0.407), maximum velocity of mitral valve A wave (p=0.050), E/A ratio (p=0.501) and tricuspid valve regurgitation pressure (p=0.668) were not different in the two groups. However, tricuspid valve regurgitation velocity was higher in diabetics (3.1 ± 0.7 vs 2.9 ± 0.5 m/s, p=0.003).

**Conclusions:** Although diabetes poses an additional burden for patients with CHF, survival is similar in diabetics and nondiabetics. Nonetheless, readmission rates may be higher among diabetics. Tricuspid return velocity is higher in diabetics, suggesting early pulmonary vasculature remodeling.

## Introduction

The prevalence of Type 2 Diabetes (T2DM) in the United States remains alarmingly high, affecting nearly 41 million individuals. Among this population, approximately 22% carry the additional diagnosis of congestive heart failure (CHF) [1,2]. T2DM is widely recognized to be a risk factor for developing CHF and is associated with increased baseline morbidity and mortality [1–4]. Large epidemiologic studies, including the Framingham Heart Study, UKPDS, and First National Health and Nutrition Examination Survey Epidemiologic Follow Up Study, have acknowledged that patients with T2DM have a two-to-four-fold increased risk of developing either systolic or diastolic CHF compared to nondiabetics [1,3]. Numerous studies have demonstrated this concept, termed “diabetic cardiomyopathy”, which refers to myocardial dysfunction attributable to diabetes itself, free of confounders such as hypertension, coronary artery disease, and ventricular heart disease [4–6]. After adjustment for other comorbidities, isolated T2DM remains an independent predictor of CHF.

The pathophysiology of T2DM leading to CHF has been studied extensively, with most authors emphasizing the importance of advanced glycation end products (AGEs) and neurohormonal dysregulation [2,4,7]. Incremental increases in hemoglobin A1c are associated with a 15-30% higher risk of incident HF, highlighting the interconnectedness of hyperglycemia and myocardial dysfunction [2,8–10]. AGEs promote arterial and myocardial stiffening via crosslinking of collagen types I and III and fibroblast stimulation [1,4,11]. At the same time, oxidative stress accelerates atherogenesis and endothelial dysfunction through modifications of low-density lipoprotein cholesterol (LDL-C) [4,12,13]. Nevertheless, the most deleterious effect of T2DM is the overactivation of the sympathetic nervous system and renin-angiotensin-aldosterone system, thereby causing cardiac remodeling and autonomic neuropathy [11] [1] [2] [14]. This chronic stimulation provides a foundation for worsening insulin resistance and oxidative stress/inflammation. To make matters worse, most patients with T2DM have additional comorbidities, which are independent risk factors for CHF, such as chronic kidney disease, coronary artery disease, hypertension, and obesity.

The increased cardiovascular risk imparted by both diabetes and CHF naturally implies that the possibility of complications is much higher compared to nondiabetics. Knowing this, it is vital to recognize the development and progression of T2DM in patients with CHF, and vice versa. Whereas most clinical practice focuses on the progression and treatment of T2DM and CHF as two separate entities, this paper aims to highlight their intimate relationship.

## Methods

We did a retrospective analysis of prospectively collected data initially reported by Zhang et al [15] using PhysioNet [16], including 2,008 patients hospitalized for CHF exacerbation between December 2016 and June 2019. A total of 166 variables were generated from the electronic data set, and blanks were used to indicate missing values. The European Society of Cardiology criteria were used to define heart failure as the presence of symptoms of heart failure, BNP >35 pg/mL or NT-proBNP >125 pg/mL and structural/functional evidence of heart failure. All types of heart failure (acute, chronic, left, right) were included. Institutional Review Board (IRB) approval was waived due to the use of unidentifiable and publicly available patient information.

On hospital admission, baseline characteristics were measured: body temperature, pulse, respiration rate (RR), blood pressure, body mass index (BMI), type of heart failure (left, right, both), and comorbidities including T2DM. Patients’ acuity was further delineated by NYHA class, need for oxygen therapy, labs, level of care, level of consciousness and Charlson Comorbidity Index (CCI). Lastly, left ventricular ejection fraction (LVEF), maximum A and E wave velocities of the mitral valve, E/A ratio, tricuspid valve regurgitation velocity and pressure were obtained from patients’ index echocardiograms.

### Outcomes

Outcome variables included discharge destination, a mandatory follow up visit at 28 days, 3 months, and 6 months, readmission within 3 and 6 months, time to death and time to readmission from index hospital admission, return to emergency department within 6 months, and death within 3 and 6 months. In addition, the echocardiographic parameters on admission of diabetics and non-diabetics were compared in a secondary analysis.

### Statistical Analysis

Propensity score matching (PSM) was used to match diabetics and nondiabetics from baseline characteristics, including demographics (age, sex, socioeconomic status), severity of CHF exacerbation (level of care, O2 support, prior CHF hospitalizations, vital signs, NYHA class, detailed laboratory findings, level of consciousness) and comorbidities. A multivariate logistic regression model was made using the presence of diabetes as the dependent variable. The probability of the dependent variable occurring was calculated for every patient. Cases were sorted based on this probability. Patients with similar probabilities among diabetics and non-diabetics were matched using the nearest neighbor method.

After PSM, outcomes in diabetics and non-diabetics were compared using univariate analysis. T-test was used for continuous variables. Chi-square and Fisher’s exact tests were used for categorical variables. The threshold of significance was set at 0.05 and two-tailed statistical tests were used for all comparisons. The analysis was done on SPSS v.22 (IBM Corporation, New York, USA).

## Results

A total of 2,008 patients were included in the study. Prior to propensity score matching, 466 patients were diabetic, among which 176 (37.8%) were males and 290 (62.2%) females. Among nondiabetics, 669 (43.4%) were male and 873 (56.6%) were female. For most patients, this was their first hospitalization for HF exacerbation (92.1% among diabetics, 92.8% among non-diabetics). According to NYHA classification, the majority of patients were diagnosed with class III disease, 225 diabetics (48.3%) and 814 non-diabetics (52.8%). Most patients had HF involving both the left and right heart, including 350 (75.1%) of diabetics and 1130 (73.3%) of non-diabetics. To reduce confounding bias, diabetic and nondiabetic patients were matched and a total of 492 patients resulted.

After PSM, 244 had diabetes (86 [35.2%] males and 160 [65.6%] females) and 248 did not (94 [37.9%] males and 152 [61.3%] females). After PSM, there were no significant differences in terms of demographics, level of care, vital signs, BMI, type of HF, NYHA classification, comorbidities, laboratory values and arterial blood gas results. Baseline patient characteristics before and after matching are described in **Table 1**.

**Table 1.** Baseline Characteristics of Diabetics and Non-Diabetics with Heart Failure Before and After Matching.

| Variables | Before Matching |  |  | After Matching |  |  |
| --- | --- | --- | --- | --- | --- | --- |
|  | Diabetics | Non-Diabetics | p-value | Diabetics | Non-Diabetics | p-value |
| Level of care |  |  | 0.760 |  |  | 0.499 |
| Regular floor | 462 (99.1%) | 1531 (99.3%) |  | 244 (99.2%) | 246 (100.0%) |  |
| Intensive Care Unit | 4 (0.9%) | 11 (0.7%) |  | 2 (0.8%) | -- |  |
| Socioeconomic status |  |  | 0.491 |  |  | 0.191 |
| Middle Class | 415 (89.1%) | 1351 (87.6%) |  | 224 (91.1%) | 215 (87.4%) |  |
| Lower Class | 46 (9.9%) | 169 (11.0%) |  | 22 (8.9%) | 31 (12.6%) |  |
| Prior visits |  |  | 0.591 |  |  | 0.553 |
| No | 429 (92.1%) | 1431 (92.8%) |  | 231 (93.9%) | 234 (95.1%) |  |
| Yes | 37 (7.9%) | 111 (7.2%) |  | 15 (6.1%) | 12 (4.9%) |  |
| Age |  |  | 0.878 |  |  | 0.834 |
| </59-69 | 128 (27.5%) | 418 (27.1%) |  | 61 (24.8%) | 59 (24.0%) |  |
| >70 | 338 (72.5%) | 1124 (72.9%) |  | 185 (75.2%) | 187 (76.0%) |  |
| Sex |  |  | 0.031 |  |  | 0.454 |
| Male | 176 (37.8%) | 669 (43.4%) |  | 86 (35.0%) | 94 (38.2%) |  |
| Female | 290 (62.2%) | 873 (56.6%) |  | 160 (65.0%) | 152 (61.8%) |  |
| Temperature (C) |  |  | 0.797 |  |  | 0.339 |
| <38 | 457 (98.1%) | 1515 (98.2%) |  | 239 (97.2%) | 243 (98.8%) |  |
| >/38 | 9 (1.9%) | 27 (1.8%) |  | 7 (2.8%) | 3 (1.2%) |  |
| Pulse (beats per minute) |  |  | 0.783 |  |  | 0.579 |
| <90 | 285 (61.2%) | 954 (61.9%) |  | 147 (59.8%) | 153 (62.2%) |  |
| >/90 | 181 (38.8%) | 588 (38.1%) |  | 99 (40.2%) | 93 (37.8%) |  |
| Respiratory Rate (breaths per minute) |  |  | 0.163 |  |  | 0.335 |
| <20 | 340 (73.0%) | 1174 (76.1%) |  | 185 (75.2%) | 194 (78.9%) |  |
| >/20 | 126 (27.0%) | 368 (23.9%) |  | 61 (24.8%) | 52 (21.1%) |  |
| Mean Arterial Pressure (mmHg) |  |  | 0.558 |  |  | 0.623 |
| <65 | 6 (1.3%) | 15 (1.0%) |  | 3 (1.2%) | 1 (0.4%) |  |
| >/65 | 460 (98.7%) | 1527 (99.0%) |  | 243 (98.8%) | 245 (99.6%) |  |
| Body Mass Index (kg/m^2) |  |  | 0.007 |  |  | 0.484 |
| <25 | 375 (80.5%) | 1320 (85.6%) |  | 204 (82.9%) | 198 (80.5%) |  |
| >/25 | 91 (19.5%) | 222 (14.4%) |  | 42 (17.1%) | 48 (19.5%) |  |
| Type of Heart Failure |  |  | 0.251 |  |  | 0.904 |
| Both | 350 (75.1%) | 1130 (73.3%) |  | 188 (76.4%) | 185 (75.3%) |  |
| Left | 109 (23.4%) | 368 (23.9%) |  | 55 (22.4%) | 57 (23.2%) |  |
| Right | 7 (1.5%) | 44 (2.9%) |  | 3 (1.2%) | 4 (1.6%) |  |
| NYHA class |  |  | 0.022 |  |  | 0.629 |
| II | 74 (15.9%) | 279 (18.1%) |  | 46 (18.7%) | 43 (17.5%) |  |
| III | 225 (48.3%) | 814 (52.8%) |  | 125 (50.8%) | 118 (48.0%) |  |
| IV | 167 (35.8%) | 449 (29.1%) |  | 75 (30.5%) | 85 (34.6%) |  |
| Comorbidities |  |  |  |  |  |  |
| Myocardial Infarction | 45 (9.7%) | 98 (6.4%) | 0.015 | 20 (8.1%) | 23 (9.3%) | 0.632 |
| Peripheral Arterial Disease | 26 (5.6%) | 75 (4.9%) | 0.536 | 15 (6.1%) | 11 (4.5%) | 0.420 |
| Cerebrovascular Disease | 37 (7.9%) | 113 (7.3%) | 0.660 | 21 (8.5%) | 27 (11.0%) | 0.362 |
| Chronic Obstructive Pulmonary Disease | 42 (9.0%) | 191 (12.4%) | 0.046 | 20 (8.1%) | 19 (7.7%) | 0.867 |
| Chronic Kidney Disease | 168 (36.1%) | 306 (19.8%) | <0.001 | 63 (25.6%) | 59 (24.0%) | 0.676 |
| CCI (Charlson Comorbidity Index Score) |  |  | <0.001 |  |  | 0.851 |
| 0-2 | 190 (40.8%) | 1335 (86.6%) |  | 158 (64.2%) | 156 (63.4%) |  |
| 3+ | 271 (58.2%) | 207 (13.4%) |  | 88 (35.8%) | 90 (36.6%) |  |
| Consciousness |  |  | 0.439 |  |  | 1.000 |
| Clear | 460 (98.7%) | 1514 (98.2%) |  | 243 (98.8%) | 242 (98.4%) |  |
| Abnormal Response | 6 (1.3%) | 28 (1.8%) |  | 3 (1.2%) | 4 (1.6%) |  |
| O2 support |  |  | 0.691 |  |  | 0.846 |
| None | 458 (98.3%) | 1508 (97.8%) |  | 243 (98.8%) | 242 (98.4%) |  |
| Non-invasive ventilation | 4 (0.9%) | 13 (0.8%) |  | 2 (0.8%) | 2 (0.8%) |  |
| Invasive mechanical ventilation | 4 (0.9%) | 21 (1.4%) |  | 1 (0.4%) | 2 (0.8%) |  |
| Acute Kidney Injury |  |  | 0.667 |  |  | --- |
| No | 464 (99.6%) | 1537 (99.7%) |  | 246 (100.0%) | 246 (100.0%) |  |
| Yes | 2 (0.4%) | 5 (0.3%) |  | - |  |  |
| White Blood Cell count |  |  | <0.001 |  |  | 0.663 |
| </11 x 10<sup>9</sup>/L | 386 (82.8%) | 1369 (88.8%) |  | 218 (88.6%) | 221 (89.8%) |  |
| >11 x 10<sup>9</sup>/L | 75 (16.1%) | 151 (9.8%) |  | 28 (11.4%) | 25 (10.2%) |  |
| Hemoglobin (g/L) |  |  | 0.004 |  |  | 0.838 |
| >100 | 324 (69.5%) | 1169 (75.8%) |  | 180 (73.2%) | 182 (74.0%) |  |
| </100 | 137 (29.4%) | 350 (22.7%) |  | 66 (26.8%) | 64 (26.0%) |  |
| Platelet count (x10<sup>9</sup>/L) |  |  | 0.168 |  |  | 0.096 |
| >/150 | 197 (42.3%) | 595 (38.6%) |  | 105 (42.7%) | 87 (35.4%) |  |
| <150 | 264 (56.7%) | 925 (60.0%) |  | 141 (57.3%) | 159 (64.6%) |  |
| INR (International Normalized Ratio) |  |  | 0.352 |  |  | 0.787 |
| </1.2 | 226 (48.5%) | 710 (46.0%) |  | 121 (49.2%) | 124 (50.4%) |  |
| >1.2 | 232 (49.8%) | 805 (52.2%) |  | 125 (50.8%) | 122 (49.6%) |  |
| High-sensitivity troponin (ng/L) |  |  | 1.000 |  |  | 1.000 |
| </14 | 450 (96.6%) | 1473 (95.5%) |  | 245 (99.6%) | 245 (99.6%) |  |
| >14 | 1 (0.2%) | 5 (0.3%) |  | 1 (0.4%) | 1 (0.4%) |  |
| Potassium<br>(mmol/L) |  |  | <0.001 |  |  | 0.343 |
| >/5 | 60 (12.9%) | 108 (7.0%) |  | 25 (10.2%) | 19 (7.7%) |  |
| <5 | 403 (86.5%) | 1426 (92.5%) |  | 221 (89.8%) | 227 (92.3%) |  |
| Sodium (mmol/L) |  |  | 0.010 |  |  | 0.991 |
| >/130-145 | 420 (90.1%) | 1410 (91.4%) |  | 228 (92.7%) | 225 (91.5%) |  |
| <130 | 35 (7.5%) | 71 (4.6%) |  | 13 (5.3%) | 15 (6.1%) |  |
| >145 | 8 (1.7%) | 53 (3.4%) |  | 5 (2.0%) | 6 (2.34%) |  |
| BNP (pg/mL) |  |  | 0.471 |  |  | 0.926 |
| <500 | 179 (38.4%) | 559 (36.3%) |  | 91 (37.0%) | 90 (36.6%) |  |
| >/500 | 282 (60.5%) | 953 (61.8%) |  | 155 (63.0%) | 156 (63.4%) |  |
| LDL (mmol/L) |  |  | 0.986 |  |  | 0.693 |
| </2.6 | 358 (76.8%) | 1188 (77.0%) |  | 211 (85.8%) | 214 (87.0%) |  |
| >2.6 | 61 (13.1%) | 203 (13.2%) |  | 35 (14.2%) | 32 (13.0%) |  |
| Creatinine<br>(umol/L) | 118.9461<br>(92.80801) | 105.9 (72.2) | <0.001 | 108.4 (75.8) | 104.3 (68.1) | 0.348 |
| Urea (mmol/L) | 10.3265<br>(5.74550) | 9.3 (5.5) | 0.012 | 9.6 (5.2) | 9.2 (5.6) | 0.817 |
| Glomerular<br>Filtration Rate<br>(mL/min/1.73 m^2) | 66.3532<br>(40.89380) | 69.4 (35.2) | 0.002 | 69.3 (39.4) | 69.4 (34.9) | 0.248 |
| APTT | 34.8020<br>(6.67085) | 35.6 (8.7) | 0.053 | 35.3 (6.8) | 36.2 (12.7) | 0.053 |
| Fibrinogen (g/L) | 3.4092<br>(1.08532) | 3.2 (1.0) | 0.225 | 3.5 (1.1) | 3.2 (1.2) | 0.264 |
| Myoglobin<br>(ng/mL) | 143.2780<br>(150.23698) | 116.6 (136.3) | 0.068 | 114.0 (102.8) | 99.1 (89.6) | 0.170 |
| Creatine Kinase<br>isoenzyme to<br>creatinine kinase | 0.2031<br>(0.15093) | 0.2 (0.2) | 0.711 | 0.20 (0.16) | 0.2 (0.2) | 0.660 |
| Hydroxybutyrate dehydrogenase to lactate dehydrogenase | 0.8043<br>(0.09919) | 0.8 (0.1) | 0.229 | 0.81 (0.1) | 0.8 (0.1) | 0.160 |
| Creatine kinase (IU/L) | 133.4010<br>(148.20639) | 137.3 (314.6) | 0.471 | 133.7 (157.8) | 149.3 (390.3) | 0.221 |
| Creatine kinase isoenzyme (IU/L) | 19.2729<br>(15.60670) | 20.2 (20.1) | 0.812 | 18.5 (14.7) | 20.8 (25.4) | 0.155 |
| Albumin (g/L) | 36.0739<br>(5.18187) | 36.7 (4.9) | 0.198 | 36.6 (5.1) | 36.4 (5.2) | 0.810 |
| Alkaline phosphatase (U/L) | 88.7234<br>(36.91613) | 89.7 (47.5) | 0.619 | 86.0 (34.0) | 91.1 (39.9) | 0.429 |
| HDL (mmol/L) | 1.0465<br>(0.34938) | 1.1 (0.4) | 0.897 | 1.1 (0.3) | 1.2 (0.4) | 0.550 |
| pH | 7.4078<br>(0.07333) | 7.4 (0.1) | 0.642 | 7.4 (0.1) | 7.4 (0.1) | 0.575 |
| Glucose blood gas | 10.3589<br>(5.71270) | 7.0 (2.8) | <0.001 | 9.9 (4.9) | 6.7 (1.7) | <0.001 |
| Bicarb (mmol/L) | 21.8272<br>(4.70911) | 22.3 (4.9) | 0.897 | 22.3 (4.7) | 23.0 (4.9) | 0.935 |
| O2 sat(%) | 95.7967<br>(3.91952) | 95.8 (6.8) | 0.212 | 95.9 (3.6) | 96.4 (4.5) | 0.642 |
| Pao2 pressure (mmHg) | 104.1504<br>(36.99859) | 109.4 (37.5) | 0.466 | 104.0 (36.3) | 111.0 (35.3) | 0.990 |
| Anion gap (mmol/L) | 14.0553<br>(3.58036) | 14.0 (4.3) | 0.190 | 13.9 (3.3) | 14.1 (4.4) | 0.106 |

NYHA: New York Heart Association Functional Classification

### Clinical Outcomes

Several outcomes were assessed, including discharge destination, mortality (at 28 d, 3 mo, 6 mo), readmissions (at 28 d, 3 mo, 6 mo) and return to the emergency department within 6 months. Discharge destination was not statistically significant between diabetics and nondiabetics (death 0.4% vs. 0.4%, healthcare facility 21.1% vs. 20.7%, home 70.3% vs. 69.5%, p=0.999). Death within 28 days (1.2% vs 2.8%, p=0.339), death within 3 months (1.6% vs 2.8%, p=0.544) and death within 6 months (2.4% vs 3.3%, p=0.588) were also not significantly different among the two groups. Readmission rates within 28 days (8.9% vs 7.7%, p=0.625) were also not different. However, there was a trend for higher readmission rates among diabetics at 3 months (29.3% vs. 21.5%, p=0.049) and 6 months (44.3% vs 35.8%, p=0.053) **(Table 2)**.

**Table 2.** Outcomes of Diabetics and Non-Diabetics with Heart Failure Before and After Matching.

| Variables | Before Matching |  |  | After Matching |  |  |
| --- | --- | --- | --- | --- | --- | --- |
|  | Diabetics | Non-Diabetics | p-value | Diabetics | Non-Diabetics | p-value |
| Discharge Destination |  |  | 0.852 |  |  | 0.999 |
| Death | 4 (9%) | 10 (0.6%) |  | 1 (0.4%) | 1 (0.4%) |  |
| Healthcare Facility | 99 (21.2%) | 339 (22.0%) |  | 52 (21.1%) | 51 (20.7%) |  |
| Home | 313 (67.2%) | 1031 (66.9%) |  | 173 (70.3%) | 171 (69.5%) |  |
| Dead within 28 days |  |  | 0.818 |  |  | 0.339 |
| No | 458 (98.3%) | 1513 (98.1%) |  | 243 (98.8%) | 239 (97.2%) |  |
| Yes | 8 (1.7%) | 29 (1.9%) |  | 3 (1.2%) | 7 (2.8%) |  |
| Readmission within 28 days |  |  | 0.003 |  |  | 0.625 |
| No | 419 (89.9%) | 1449 (94.0%) |  | 224 (91.1%) | 227 (92.3%) |  |
| Yes | 47 (10.1%) | 93 (6.0%) |  | 22 (8.9%) | 19 (7.7%) |  |
| Dead within 3 months |  |  | 0.926 |  |  | 0.544 |
| No | 456 (97.9%) | 1510 (97.9%) |  | 242 (98.4%) | 239 (97.2%) |  |
| Yes | 10 (2.1%) | 32 (2.1%) |  | 4 (1.6%) | 7 (2.8%) |  |
| Readmission within 3 months |  |  | 0.001 |  |  | 0.049 |
| No | 323 (69.3%) | 1187 (77.0%) |  | 174 (70.7%) | 193 (78.5%) |  |
| Yes | 143 (30.7%) | 355 (23.0%) |  | 72 (29.3%) | 53 (21.5%) |  |
| Death within 6 months |  |  | 0.942 |  |  | 0.588 |
| No | 453 (97.2%) | 1498 (97.1%) |  | 240 (97.6%) | 238 (96.7%) |  |
| Yes | 13 (2.8%) | 44 (2.9%) |  | 6 (2.4%) | 8 (3.3%) |  |
| Readmission within 6 months |  |  | <0.001 |  |  | 0.053 |
| No | 253 (54.3%) | 982 (63.7%) |  | 137 (55.7%) | 158 (64.2%) |  |
| Yes | 213 (45.7%) | 560 (36.3%) |  | 109 (44.3%) | 88 (35.8%) |  |
| Return to<br>Emergency<br>Department<br>within 6 months |  |  | 0.001 |  |  | 0.053 |
| No | 254 (54.5%) | 978 (63.4%) |  | 137 (55.7%) | 158 (64.2%) |  |
| Yes | 212 (45.5%) | 563 (36.5%) |  | 109 (44.3%) | 88 (35.8%) |  |

### Echocardiographic characteristics

Echocardiographic characteristics, including LVEF (52.2% vs 51.8%, p=0.135), LV EDV (10.4 cm vs 51.6 cm, p=0.707), maximum velocity of mitral valve E wave (7.7 m/s vs 10.m/s, p=0.407), maximum velocity of mitral valve A wave (6.4 m/s vs 0.8 m/s, p=0.050), E/A ratio (1.2 vs 1.3, p=0.501) and tricuspid valve regurgitation pressure (33.2 mmHg vs 33.4 mmHg, p=0.668) were not different in the two groups. However, tricuspid valve regurgitation velocity was found to be higher in diabetics (3.1 ± 0.7 vs 2.9 ± 0.5 m/s, p=0.003) **(Table 3)**.

**Table 3.** Echocardiographic Findings.

|  | Diabetics | Non-Diabetics | p-value |
| --- | --- | --- | --- |
| LVEF (%) | 52.2 (13.9) | 51.8 (12.3) | 0.135 |
| LV EDV (cm) | 53.0 (10.4) | 51.6 (10.4) | 0.707 |
| Maximum Velocity of Mitral Valve E Wave (m/s) | 7.7 (51.6) | 10.7 (62.0) | 0.407 |
| Maximum Velocity of Mitral Valve A Wave (m/s) | 6.4 (45.2) | 0.8 (0.3) | 0.050 |
| E/A ratio | 1.2 (0.7) | 1.3 (0.7) | 0.501 |
| Tricuspid Valve Regurgitation Velocity (m/s) | 3.1 (0.7) | 2.9 (0.5) | 0.003 |
| Tricuspid Valve Regurgitation Pressure Gradient (mmHg) | 33.2 (12.6) | 33.4 (10.9) | 0.668 |

LVEF: Left Ventricular Ejection Fraction, LV EDV: Left Ventricular End Diastolic Volume

## Discussion

Given the rising prevalence of both T2DM and CHF in the United States, it is important to understand how T2DM influences the clinical trajectory and outcomes of CHF. In our study, mortality was not different between diabetics and non-diabetics, but there was a trend for higher readmissions among diabetics. To our knowledge, this is the first head-to-head comparison of diabetics and non-diabetics in patients with CHF.

Various studies have shown that T2DM itself is an independent risk factor for worse outcomes in patients with CHF [2,3,8,11,17–19]. For example, in post-hoc analyses of the Candesartan in Heart failure: Assessment of Reduction in Mortality and morbidity (CHARM) program, diabetes mellitus was an independent predictor of all-cause mortality and the composite endpoint of cardiovascular death or heart failure hospitalization [20–22]. Another study by Dauriz et al found that DM was “diabetes was associated with a 28% increased risk of all-cause mortality and approximately 35% risk of both CV death and hospitalization (mostly from HF) over a three year follow up [23]. Similarly, the Atherosclerosis Risk in Communities study illustrated that elevated A1c is a risk factor for development of CHF [10]. In this study, we similarly show that while DM was not associated with a higher mortality rate, readmissions were higher among diabetics. While this study may be underpowered to detect differences in mortality, it demonstrated that in a head-to-head comparison, DM is associated with higher readmission rates.

In other studies, T2DM has been found to be a risk factor for death and readmission in CHF. For example, The ESC-HFA long-Term Registry highlighted that T2DM independently predicts death and rehospitalization after acute heart failure, and Kong et al further delineated the increased mortality seen in acute HFrEF exacerbations specifically [24,25]. Agarwal et al showed higher death rates among diabetics compared to non-diabetics with either HFrEF or HFmREF [26]. Chronic heart failure was studied in the SOLVD Trials and Registry, CHARM trial, and I-Preserve Diabetes Analysis, each of which showed worse mortality from HFrEF in diabetics vs nondiabetics [20–22,27,28]. Additionally, the UK Prospective Diabetes Study emphasized that T2DM carries a three-fold increased risk for all cardiovascular diseases, affecting morbidity across multiple pathologies [29].

Our findings suggest that, in the context of contemporary heart-failure care, diabetes may preferentially contribute to recurrent congestion and hospitalization rather than excess short-term mortality. This pattern aligns with prior observations in heterogeneous heart-failure populations, particularly in HFpEF, where diabetes is more consistently associated with increased morbidity rather than excess mortality [25,28,30–32], especially after adjusting for confounders. Similar to our analysis, literature regarding HFpEF have shown more mixed results regarding mortality. Although T2DM in HFpEF is associated with worsened diastolic function, higher comorbidity burden, and increased hospitalizations, the association with mortality is more attenuated after adjustment compared to HFrEF [2,20,22,33–35]. For example, Echouffo-Tcheugui et al [31] illustrated that there is no independent association with in-hospital mortality for acute HFpEF exacerbation after multivariate analysis. Likewise, the analysis of the I-PRESERVE study found that diabetes is associated with increased HF hospitalizations and worse functional status, though the mortality signal after adjustment was more attenuated and sensitive to factors such as renal function and age [36]. In the TOPCAT post-hoc analyses, patients with T2DM and HFpEF have more significant volume overload, higher BNP levels, and greater burden of structural/functional echocardiographic changes, but not necessarily worse outcomes in regard to mortality [37]. Multiple other cohorts have demonstrated this, especially in the acute populations [25,26,32,38].

In terms of echocardiographic findings, several markers of LV systolic and diastolic function were not found to be statistically significant between the two groups, including LVEF, LV EDV, Maximum velocity of the mitral valve A wave and E wave, and the E/A ratio. Interestingly, the Tricuspid Regurgitation Velocity (TRG) was statistically significant in diabetics vs nondiabetics, though TRPG was not. Knowing that TRV is an index for higher RV pressure, and thus higher pulmonary pressure, TRV is commonly used to assess for elevated left-sided pressures and secondary pulmonary hypertension [39]. While TRV is a measured value, TRPG is calculated using Bernoulli’s equation. It reflects the pressure gradient between the right atrium and right ventricle during systole. Although both are used to screen for pulmonary hypertension, the fact that TRPG is a calculated value “magnifies variability”, and thus statistical significance. TRPG is calculated by 4 x (TRV)^2^, and when values are squared, confidence intervals widen, and p-values can become nonsignificant. Since TRV is not squared, the distribution is much tighter and therefore has more statistical power. Additionally, since TRPG depends on TRV, errors in the measurement of TRV amplify in TRPG. Given this, our paper will report TRV as the primary variable when assessing for the screening of pulmonary hypertension. Although both diabetics and nondiabetics with heart failure are at higher risk for increased LV stiffness and left sided pressures, diabetic patients have additional contributors. For example, the microvascular and endothelial dysfunction caused by hyperglycemia augments impaired pulmonary vasodilation, increased stiffness of vessels, and causes higher pulmonary vascular resistance. Patients with both T2DM and heart failure are also more prone to obesity, which leads to higher filling pressures due to increased systemic inflammation leading to myocardial fibrosis, larger total blood volume (preload) and sodium retention which activates RAAS, and renal dysfunction which leads to volume overload. The fact that TRPG was not statistically significant could also illustrate early disease that is not severe enough to produce a meaningful TRPG separation.

The study has several limitations to be discussed. First, it is subject to residual confounding given its retrospective nature. Additionally, diabetes status was indicated by yes or no, and did not capture the true severity of the individual’s disease, namely by duration, A1c, insulin use, or microvascular complications. This therefore can dilute the significance of the associations with CHF. Lastly, CHF was classified by left or right, not by HFpEF or HFreF. Knowing whether the patient suffered from diastolic vs systolic vs mixed disease would allow for more specificity regarding the effects of T2DM.

## Conclusions

This paper aims to highlight the pathophysiology and features shared between T2DM and CHF as well as emphasize the effect on outcomes and the importance of early treatment. Within this dataset, the presence of T2DM demonstrated a trend towards increased readmission rates. After adjustment for baseline characteristics and treatment received, T2DM was not independently associated with increased all-cause mortality. Therefore, while T2DM may intensify the burden of heart failure–related hospitalization, its impact on long-term survival may be reduced given the advances in heart failure management. Further studies are needed to determine whether specific phenotypes of heart failure, A1c-lowering strategies, or cardiometabolic therapies modify risk in diabetic versus non-diabetic populations.

## Data Availability

All data are available online at https://physionet.org/content/heart-failure-zigong/1.3/

